# Risk of Stress/Depression and Functional Impairment in Denmark Immediately Following a COVID-19 Shutdown

**DOI:** 10.1101/2020.12.15.20248251

**Authors:** Lars H. Andersen, Peter Fallesen, Tim A. Bruckner

## Abstract

**Background:** This study aimed to investigate the impact of the first COVID-19 lockdown (March-April 2020) on risk for stress/depression and functional impairment in a representative sample of adult individuals in Denmark, and whether the impact of lockdown was heterogeneous across living situation.

**Methods:** Using a representative, randomly drawn sample from the complete Danish adult population interviewed in March 2 to April 13, 2020 (n=2,836) and again in July 2020 (n=1,526, 54% retention rate), we study how the imposed lockdown announced March 11 following the onset of the first Danish wave of COVID-19 infections affected mental wellbeing. We use the World Health Organization Five Well-being Index (WHO-5) and the Work and Social Adjustment Scale (WSAS) to capture wellbeing and functioning. Using covariate adjusted ordinary least squares linear probability models and exploiting variation in the timing of responses occurring just before and just after the introduction of lockdown, we compare respondents before lockdown to respondents that answered during lockdown, as well to answers in re-interviews in July.

**Results:** We find reduced depressive symptoms among adults immediately after the shutdown, concentrated in adults with children living at home. Measures of functional impairment also decline immediately after the March shutdown among adults with children living at home. Impairment intensified for the entire sample between March and July, but depressive symptoms remained at lower rate in July.

**Conclusions:** Findings in Denmark indicate that living with children at home may have, in the short term, buffered the potential mental health sequelae of the COVID-19 shutdown.

## INTRODUCTION

As of mid-October 2020, more than 90 countries across the world imposed some form of lockdown in the wake of the COVID-19 pandemic [1]. Lockdowns range in scope and duration, but all imply a degree of social isolation as well as disruption from routine social, educational, and/or work activity. Previous research which predates COVID-19 indicates that, following social isolation and disruptions from work routines, mental wellbeing may decline [2–5], and whether people live together with others or not can be an important stratifying factor [6]. In addition, a review of mental health following more extreme measures of quarantine finds long-term psychological sequelae [7]. Taken together, this literature has raised the concern of a “second pandemic” of morbidity due to mental health problems following COVID-19 [8].

Recent research examines mental wellbeing following COVID-19 and the associated lockdowns. Three population-representative studies—in the UK, the US, and France— appear in the literature. All studies report worse mental health in Spring 2020 relative to previous years [8–11]. Ettman and colleagues, for instance, find a much greater prevalence of depressive symptoms among adults in the US in April 2020 relative to 2017/2018.

This work, while important, has two key limitations. First, the UK, the US, and France all rank in the top 15 worldwide in COVID-19 deaths per population as of November 30, 2020 [12]. This circumstance leaves open the question of whether experienced national severity of the pandemic (through media reports [8] or through direct experiences), or the social and work restrictions imposed by a lockdown *per se*, drive results. Second, none of these studies includes measures of mental health and/or wellbeing immediately *before* the lockdown. The absence of “baseline” mental health information in weeks before the lockdown raises the concern of confounding by trends over time in mental health that coincide with, but are not caused by, the COVID-19 lockdown. Third, unlike for previously studied countries, the Danish lockdown was imposed uniformly and rapidly following the first infections and came into effect before the first Danish registered COVID-19 fatality (see Figure 1).

**Figure 1.**
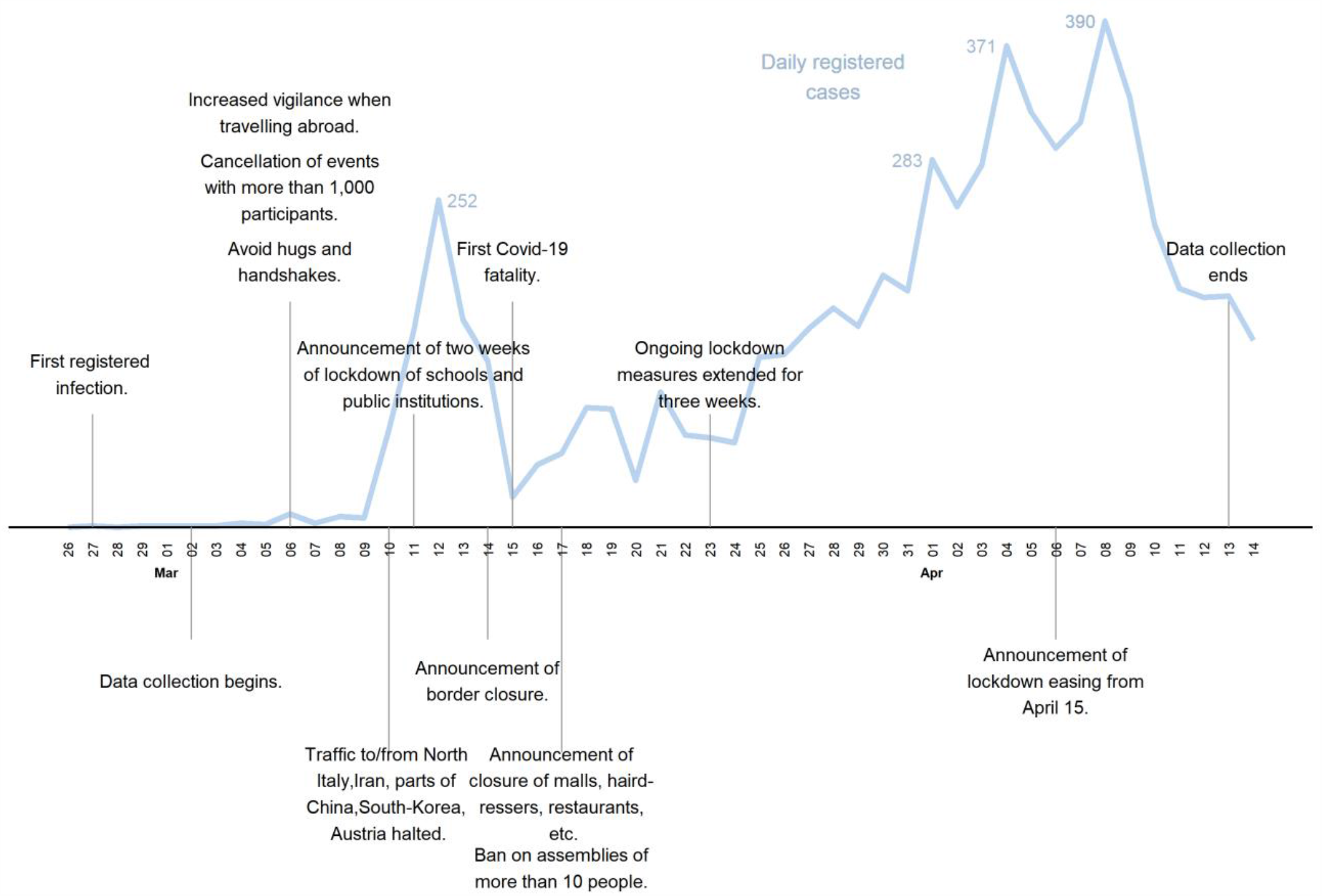
COVID-19 and lockdown development during first wave of data collection, February-April 2020.

We address these limitations and extend prior work by examining mental wellbeing in Denmark, a country that imposed a lockdown in March 2020 but reports a substantially lower COVID-19 burden (i.e., 14.3 deaths per 100,000 population) than does France, the US, or the UK (i.e., 78 to 87 deaths per 100,000 population) [12]. We also exploit variation in the timing of responses to a nationally representative survey collected in March 2020. On March 11, Denmark imposed nationwide school closures and the closing of public institutions. Survey responses occurred immediately before and after the date when the first COVID-19 lockdown was ordered and imposed.

We measured mental wellbeing among the adult Danish population through the World Health Organization Five Well-being Index (WHO-5) and the Work and Social Adjustment Scale (WSAS). These scales capture both pre-clinical measures of mental disorder as well as impairment. Further, we re-interviewed the sample in July 2020 when COVID-19 precautions were substantially lessened compared to the lockdown period in March. Given that previous research on some subgroups finds *improved* wellbeing following COVID-19 [13], we specified all tests as two-tailed. We, moreover, explored the relation between the lockdown and mental wellbeing by family structure, given that state-imposed limitations on social activity may affect persons living alone differently than for persons living with family members as suggested by prior research.

## BACKGROUND

### Lockdown timeline for Denmark

Figure 1 provides a timeline of the Danish COVID-19-restrictions, the number of confirmed cases for March and start of April 2020, and the data collection window for the first wave of the survey [1, 14]. Denmark reported its first confirmed SARS-CoV-2 case on February 27, 2020 [1]. On March 11, Denmark initiated nationwide school closures and the closing of public institutions, as the cumulative number of confirmed infections had increased to 264. Lockdown measures were further strengthened over the following six days to include border closures, the closure of restaurants, malls and hairdressers, and general encouragements to work from home. Financial aid packages to businesses and furloughed employees were launched in the same six-day period. Unlike other European countries, Denmark did not introduce curfews, stay-home orders, or mandatory use of masks during the Spring lockdown. The Danish government began easing lockdown measures from April 15, 2020. Lockdown measures were continuously eased over the summer, and not re-introduced until September 2020. Based on this timeline, we defined the beginning of the lockdown measures as March 11, 2020.

## METHODS

### Variables and Data

To consider how measures of wellbeing and impairment changed following the imposed lockdown, we used a representative survey carried out by Statistics Denmark on behalf of the Capital Region of Denmark’s Mental Health Services during March-April 2020. Using a random draw from the present population database of all Danish residents aged 18 and above, Statistics Denmark initially contacted 8,300 people through personal digital postboxes that are linked directly to people’s unique social security numbers and used for communications between Danish residents and governmental institutions. Respondents answered through computer assisted web interviews (CAWI), with those initially failing to respond receiving prompts by message to their digital postbox. The response rate to the first wave of the survey was 34 percent (N=2,836); 1,127 respondents completed the survey prior to the lockdown announcement and initiation on March 11, and 1,709 respondents completed it after March 11. These numbers reflect respondents who provided valid responses to all items of our dependent variables. Respondents who completed the survey before and after March 11 were generally alike across the background characteristics, although the proportion of respondents age 60+ decreased and the proportion of respondents with children living at home increased slightly (see Appendix Table A1).

With permission from the Capital Region of Denmark’s Mental Health Services, we then carried out a follow up survey in July 2020, where the same respondents were re-interviewed. Of respondents participating in the first wave, 1,526 (54%) also participated in the second wave collected in July 2020. Younger respondents and respondents with children living at home were less likely to participate in the second wave, as were respondents who experienced significant functional impairment in early March (which will likely cause us to underestimate a potential increase in functional impairment from March to July), but respondents in the second wave generally resemble respondents in early March (see Appendix Table A1). Answers to the survey can be linked at the individual level with administrative data from Statistics Denmark. In addition to survey data on mental wellbeing and functioning, our data therefore contain information on age, gender, living arrangement (single/in a relationship), whether respondents were living with any children in the home, region of residents (Nomenclature of Territorial Units for Statistics, level 2 ([NUTS-2]), and employment status (employed, unemployed, outside the labor force).

To capture wellbeing and experienced functional impairment, the survey included two validated measures—the WHO-5 and the WSAS—that both have distinct clinically relevant threshold values. The WHO-5 is a sensitive and specific clinical screening tool for risk of stress and depression that uses five items to capture risk of depression measured between 0-100, with each scale contributing 0-20 points. For this measure, 0 indicates the most severe depressive symptoms and 100 indicates no symptoms. The WHO-5 has strong construct validity as a unidimensional scale [15]. For Denmark, the population norm is established as 70 for adults [16]. For the WHO-5 index, we (consistent with work in clinical settings) used the established cutoff point at 50 to create a binary category for whether a respondent is at risk for depression and stress (i.e. WHO-5 < 50, [15, Table 2]).

The WSAS is a functional impairment measure designed to measure a patient’s perceived functional impairment following health problems across five items [17]. Although not originally intended for non-clinical populations, the WSAS displays valid psychometric properties across different patient populations that cover both mild and more severe (psycho-)somatic [18–20] and mental health [17, 21–23] conditions.

Furthermore, the WSAS captures a dimension of impairment distinct from depression [23]. Although not validated in a Danish version, it has previously been used both in Danish clinical and research settings [21]. WSAS measures impairment on a scale from 0 (no impairment) to 40 (most severe impairment). Given that we study a non-clinical sample, we use the cutoff point at a WSAS score of 10, with any score above 10 indicating at least significant functional impairment with or without additional psychopathologies (WSAS > 10 [17]).

### Analytical strategy

Our analytical strategy exploits the fact that data collection took place across the announcement and initiation of lockdown in Denmark in March 2020. First, we use ordinary least squares linear probability models to compare the outcomes between respondents who answered the survey before and after lockdown began. We adjust all models for gender and age. Next, in fully adjusted regression models, we control for NUTS2-region of residence, labor market status (employed, unemployed, outside the labor force), relationship status (single, married/cohabiting), and whether respondents had children at home.

The impact of lockdown could differ according to the home environment. Persons living with family, for instance, may experience relative more social interaction than would persons not living with family during the imposed lockdown. To explore this possibility, we performed sub-sample analyses that compares single individuals to individuals who are living with a partner, as well as sub-sample analyses that compare people living without children in the home to people living with children in the home, and test for differences between subgroups using a Chow-test.

In addition, to fully leverage our data structure with re-interviews in July 2020, we then compared the reported levels of risk of depression and stress and significant functional impairment measured prior to lockdown in March to the levels experienced by the same persons in July accounting for repeated measures of the same individuals with clustered standard errors. The latter exercise captures the development in the outcomes across the first wave of COVID-19 in Denmark. Here, we control for relatively few variables in the regression because we use within-individual variation across survey waves (e.g., the age of the individual does not change substantially from March to July). All calculations were carried out using Stata 15/MP.

We then performed several robustness checks. First, we evaluated whether our choice of thresholds in the outcome variables (WHO-5 < 50 and WSAS > 10) affect inference. Second, because our sample is not fully identical to the Danish population on characteristics such as gender and age, we replicate main results using population weights provided by Statistics Denmark instead of controlling for covariates. Third, selective survey participation across the lockdown in March and across the two survey waves (if, for example, people who experience increased mental distress due to the lockdown are less likely to participate than before the lockdown or they are more likely to not respond to survey wave 2) could invalidate our results (Appendix Table A1 showed some sign of such selection from wave 1 to 2, although not to any discernable degree from early to late March, on observed characteristics). To address this, we use the within person changes in response between March and July. As there was very little change in lockdown measures in July, all returning respondents recompleted the survey under identical lockdown circumstances. If our main pattern of results between respondents who answered prior to and during lockdown in March persist once we take into account individual change up to the post-lockdown July wave, it would indicate results are robust to differential selection in the first wave of response across the lockdown period, and results would thus at least be internally valid. Fourth, the items of the WHO-5 ask respondents to consider their experiences during the preceding two weeks. To account for the possibility that respondents answering within two weeks after lockdown have to consider both time before and after lockdown, as a robustness check we weighted answers given in the two weeks after lockdown with the amount of time since the lockdown announcement. If people considered a full two-week horizon it would mean that our main estimates of the impact of the lockdown order in March will be biased toward zero—that is, our main estimates would be conservative. Last, because our main results focus on respondents with children living at home, and because there may be different requirements and worries associated with having children at different ages at home, we checked whether results differ by age of the children (which we obtained from the general registers).

## RESULTS

Table 1 describes the sociodemographic characteristics of the survey participants and the population. Respondents (n=2,836) are similar to the broader Danish adult population in terms of geographical region and socioeconomic status. We observe some dissimilarities for gender and age, which we therefore control for in all reported results. Over half (54%) of participants in the first survey wave in March 2020 completed the survey in July 2020 (Appendix Table A1). In addition, during the first survey wave, 40% completed the survey before the lockdown (March 11^th^), and 60% completed the survey after the lockdown, which permits adequate sample size to estimate mean levels of depressive symptoms and functional impairment during these two distinct periods in March.

**Table 1.** Descriptive statistics of sociodemographic characteristics of survey participants and the population of 18-79-year-old people in Denmark.

| Variable | Survey |  | Population |  | T-test | p-value |
| --- | --- | --- | --- | --- | --- | --- |
|  | M | SD | M | SD |  |  |
| Male | 0.449 | 0.497 | 0.500 | 0.500 | -5.399 | <0.001 |
| Female | 0.551 | 0.497 | 0.500 | 0.500 | 5.399 | <0.001 |
| Ages 18-29 | 0.138 | 0.345 | 0.205 | 0.404 | -8.839 | <0.001 |
| Ages 30-39 | 0.109 | 0.312 | 0.155 | 0.362 | -6.835 | <0.001 |
| Ages 40-49 | 0.157 | 0.364 | 0.172 | 0.378 | -2.112 | 0.035 |
| Ages 50-59 | 0.226 | 0.418 | 0.183 | 0.387 | 5.844 | <0.001 |
| Ages 60-69 | 0.205 | 0.404 | 0.153 | 0.360 | 7.715 | <0.001 |
| Ages 70-79 | 0.165 | 0.371 | 0.131 | 0.337 | 5.356 | <0.001 |
| Singles | 0.301 | 0.459 | 0.353 | 0.478 | -5.714 | <0.001 |
| Couples | 0.699 | 0.459 | 0.647 | 0.478 | 5.714 | <0.001 |
| No children at home | 0.675 | 0.469 | 0.632 | 0.482 | 4.727 | <0.001 |
| Children at home | 0.325 | 0.469 | 0.368 | 0.482 | -4.727 | <0.001 |
| No. children age 0-2 | 0.042 | 0.253 | 0.054 | 0.234 | -2.667 | 0.008 |
| No. children age 3-5 | 0.070 | 0.311 | 0.078 | 0.291 | -1.351 | 0.177 |
| No. children age 6+ | 0.397 | 0.780 | 0.439 | 0.807 | -2.749 | 0.006 |
| Northern Jutland | 0.104 | 0.305 | 0.102 | 0.303 | 0.311 | 0.756 |
| Central Jutland | 0.245 | 0.430 | 0.227 | 0.419 | 2.278 | 0.023 |
| Southern Denmark | 0.208 | 0.406 | 0.209 | 0.407 | -0.096 | 0.924 |
| Capitol | 0.295 | 0.456 | 0.318 | 0.466 | -2.642 | 0.008 |
| Zealand | 0.148 | 0.356 | 0.144 | 0.351 | 0.629 | 0.529 |
| In Job | 0.652 | 0.476 | 0.643 | 0.479 | 1.069 | 0.285 |
| Unemployed | 0.019 | 0.138 | 0.021 | 0.144 | -0.702 | 0.483 |
| Outside labor force | 0.324 | 0.468 | 0.335 | 0.472 | -1.251 | 0.211 |
| N | 2,836 |  | 4,359,539 |  |  |  |

Note: No. children (top coded at three children) and labor market status (In Job, Unemployment, and Outside labor force) are measured from the general registers during the latest available data year (2019).

Bivariate comparison of our outcomes across the lockdown do not reach conventional levels of statistical detection. Still, adults interviewed after the lockdown in March have a slightly lower prevalence of depression and stress when compared to those interviewed before the lockdown (i.e., 20% vs. 22%, see Appendix Table A1), and this finding is statistically detectable when controlling for age and gender (column 1 of Table A2) yet does not reject the null in the fully adjusted model that controls for additional individual covariates, such as household structure (to which we return; see Appendix Table A2, column 2). If we expand the data to include respondents with valid responses to the WHO-5 items but who had not responded to the WSAS, the decline becomes statistically detectable (i.e., 20% after the lockdown and 23% before, p<.05, N=3,110). Functional impairment scores from the WSAS also show a slight decline in late March relative to pre-lockdown (i.e., 17% vs. 16%, see Appendix Table A1), but this difference does not reach conventional levels of statistical detection when we control for age and gender (Appendix Tables A1 and A2).

Following the state-imposed limitations on social activity, adults may have relied more on family members for social interaction than they did before the lockdown. Persons living alone, however, may have experienced fewer interactions during the lockdown, which implies the possibility of heterogeneous impact of the lockdown orders. We therefore classified the sample by cohabitation status, and then by whether the adult respondent reported children living at home. Of these subgroups, only adults with children living at home (upper panel of Figure 2) show a lower prevalence of depressive symptoms in late March (i.e., 17% vs. 32% in early March; p<.01, see Table A3 for adjusted regression results). Using a Chow-test, we found that the decrease for adults with children in the home compared to adults without children in the home is larger to statistically detectable degree (p <.05). All other subgroups report no difference in depressive symptoms between early to late March.

**Figure 2.**
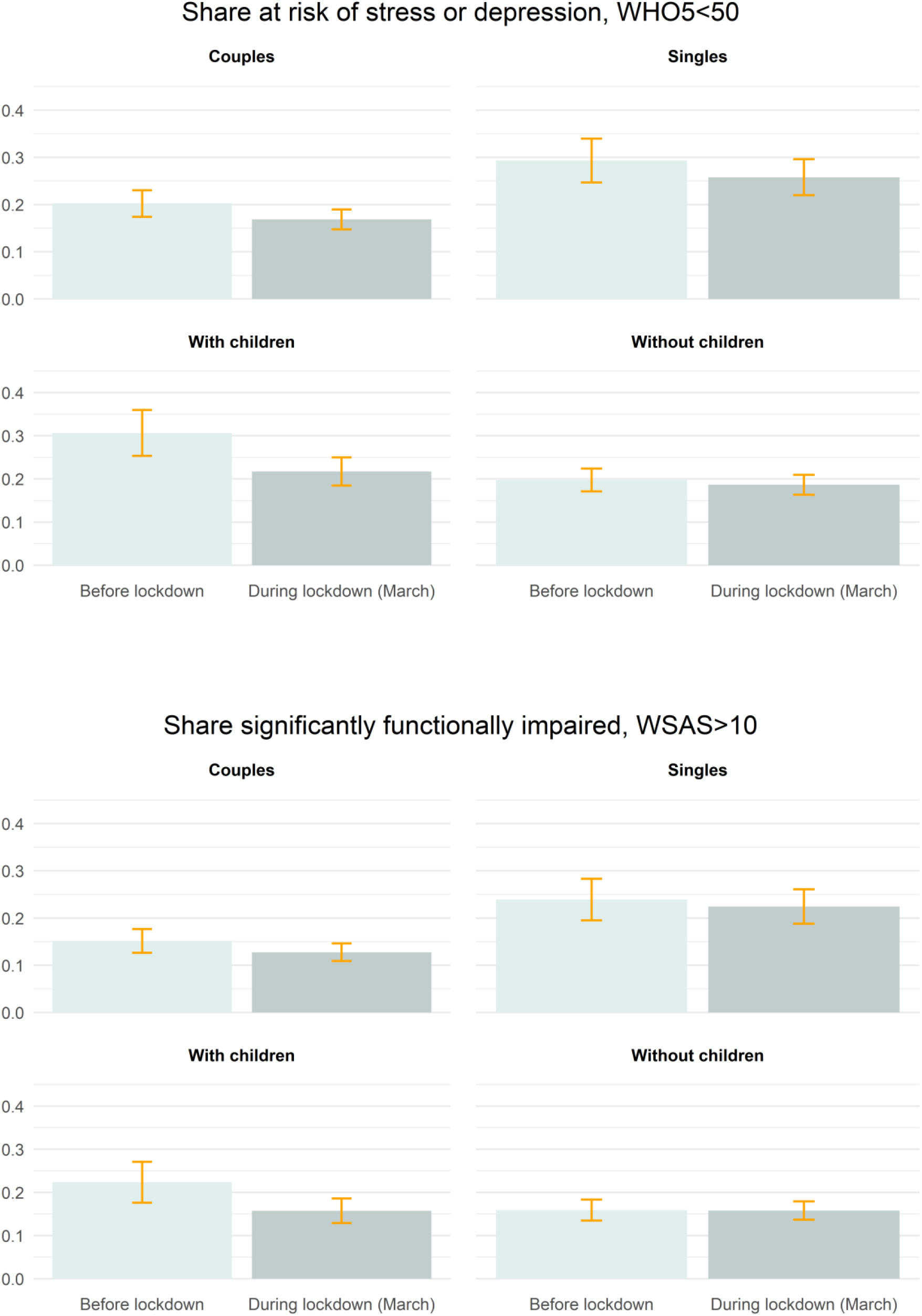
Proportion of respondents at risk of depression or stress according to the WHO5 and experiencing significant functional impairment according to the WSAS, by time of completing the survey relative to lockdown and by household structure.

The time course of WSAS functional impairment scores largely coheres with that of the subgroup trends for depressive symptoms. Adults with children living at home show a lower prevalence in functional impairment in late, relative to early, March (lower panel of Figure 2; p<.05, see Appendix Table A4). Again, using a Chow-test, we found that the decrease for adults with children in the home compared to adults without children in the home is larger to statistically detectable degree (p <.05). The lower prevalence of functional impairment among adults with children living at home remains relatively constant across WSAS sub-domains of work, social, and home functioning (Appendix Figure A1). We, by contrast, find no difference in functional impairment scores among other subgroups when comparing pre-vs. post-lockdown periods in March (lower panel of Figure 2 and Appendix Table A4).

Lockdown restrictions eased on April 15^th^. We examined whether depressive symptoms and functional impairment differed among respondents several months later—arguably once COVID-related social, economic, and institutional conventions in Denmark stabilized for a while. We restricted the study sample to persons who completed the survey in early March (i.e., pre-lockdown) and again in July 2020 (Appendix Figures A2-A4 show results including late March respondents). In aggregate, depressive symptoms among these adults fell, but functional impairment rose, in July relative to early March (p<.05 for both tests—see Appendix Table A5).

When disaggregating the July responses by family structure, only adults with children living at home and adults in couples reported a reduction in depressive symptoms in July relative to early March (upper panel of Figure 3; p<.05, see Appendix Table A6). By contrast, adults with no children at home as well as singles show no change in depressive symptoms over time (p=.713 and p=.081, respectively, see Appendix Table A6). Functional impairment, however, increased in July for all groups, albeit to a much greater extent for adults without children living at home (i.e., 13% to 35% in July; see lower panel of Figure 3 and Appendix Table A7). The increase in functional impairment in July among adults with children living at home was much lower (but still statistically significant, p<.05).

**Figure 3.**
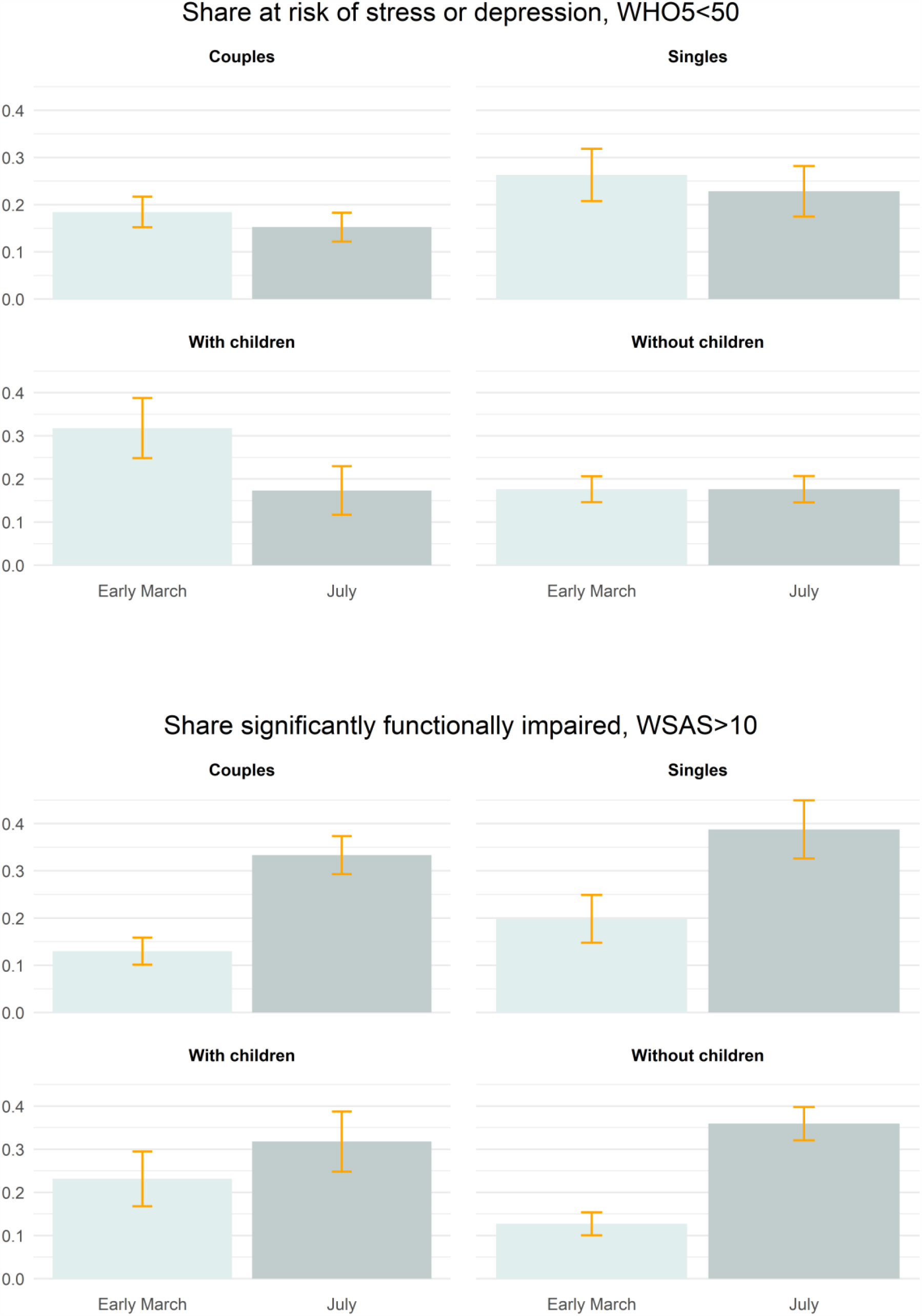
Proportion of respondents at risk of depression or stress according to the WHO5, repeat responses in early March and July, by household structure.

The results from our robustness checks do not raise concern over the validity of our main results. First, modifying the thresholds used to define depressive symptoms and functional impairment did not substantially change results (see Appendix Table A8). Second, using statistical weights provided by Statistics Denmark instead of controlling for covariates did not affect inference (see Appendix Figure A5). Third, relying on within-individual differences in the outcomes pre- and post-lockdown in March compared to July answers did not change main results (see Appendix Table A9). Fourth, down-weighting “exposure” to the lockdown among respondents who participated on March 12^th^ to 25^th^, as described in the Methods section, did not affect inference (see Appendix Figure A6; we ran the same robustness check for the WSAS and again found results similar to the main Tables (Appendix Figure A7). Results from our last robustness check (focusing on age of children) shows that our main results for respondents with children living at home are robust across age of the children when focusing on the risk of depression or stress, but that the early to late March decrease in the proportion experiencing significant functional impairment is driven by respondents with children older than 6 years (see Appendix Figures A8 and A9).

## DISCUSSION

We exploit the unique timing of a population-based behavioral survey to examine whether a COVID-19 related shutdown preceded an acute change in depressive symptoms and functional impairment. We examined these responses in Denmark, a country that experienced relatively low incidence of SARS-CoV-2 infections, but which instituted strong lockdown restrictions in March 2020. Contrary to reports in other countries, we find *reduced* depressive symptoms among adults immediately after the shutdown. This reduction, moreover, concentrates in adults with children living at home. Measures of functional impairment also decline immediately after the March shutdown, but only among adults with children living at home. Findings in Denmark indicate that living with children at home may have, in the short term, buffered the potential mental health sequelae of the COVID-19 shutdown. If others replicate our work, strengthening the type of social support that already seems to be present in families may serve as one potential avenue for minimizing the mental health sequelae of extended COVID-19 shutdowns.

Raabe and colleagues’ survey of scientists in three European countries coheres with our findings in that they report improved wellbeing immediately after the COVID-19 lockdown [13]. Similarly, Mari and colleagues find results that mirror ours across residential patterns, although they are limited to studying Italians during lockdown [11]. In contrast, a multinational study using data collected late March to early April 2020 generally find that families report the most stress during lockdown [24], but these results may simply reflect differences already existing prior to the pandemic (as our results also suggest). Whereas we hesitate to draw population-based lessons from this select survey of well-educated scientists, the authors note that strong security of employment may have contributed to their short-term satisfaction with a slower pace and a flexible work-life organization. This financial security may be similar to the situation of most Danish households during the COVID-19 pandemic. Furthermore, social cohesion may increase following adverse events given that shared adversity can connect individuals to a broader goal and purpose than before the event [25, 26]. This social cohesion explanation seems consistent with reports of fewer than expected suicide deaths immediately following the first set of COVID-19 restrictions in Germany and Japan [27, 28]. We note, however, that this explanation is necessarily *post hoc* and requires further refinement and testing before being considered as anything other than informed speculation. We also point out that the reductions in depressive symptoms among Danes appear confined to adults living with children.

Whereas adults living with children show reduced depressive symptoms in July (relative to pre-shutdown), they are more likely to report significant functional impairment in July. We suspect that, as they habituate to the reality of a prolonged COVID-19 pandemic, the ability to flexibly balance work, family, and social expectations may become strained. Interestingly, of any subgroup, adults living with children show the lowest rise in significant functional impairment in July 2020. This result should encourage further investigation, in both Denmark and elsewhere, of elements of family life that may benefit social connectivity and general mental health functioning during COVID-19.

Strengths of our study include the population-based nature of the survey, the use of two different measures of mental wellbeing and functioning, and the fact that survey responses fall immediately before and after the announced lockdown. Limitations involve the fact that the March comparisons of mental wellbeing before and after the lockdown examine serial cross-sections rather than a panel. We, however, controlled for compositional changes of the panel in our analyses. The WHO-5 also asks about 14-day recall of depressive symptoms, which may have biased pre-vs. post March 11 responses towards the null. We, however, controlled for this circumstance using a weighted analysis as a robustness check; findings, moreover, rejected the null, which precludes a type II error. Lastly, we cannot rule out the possibility of seasonal confounding in that late-March coincided with Spring and better temperature than in early March. This seasonal confounding, however, cannot explain the distinct nature of the subgroup responses in which depressive symptoms and functional impairment fall in late-March only among adults with children but not among adults living alone.

Our findings diverge from previous population-based reports in the UK, the US and France. This circumstance could arise for several reasons. First, Denmark underwent a much less severe COVID-19 pandemic in Spring 2020 than did these countries, as measured by overall cases or deaths per population. Danes, therefore, may not have had to contend as heavily with the associated fear and anxiety of COVID-19-related morbidity as did other countries. Second, Denmark’s strong social safety net largely protects adults and families against large financial “shocks” that appear more common in other countries (e.g., the US) when adults lose jobs [29]. Third, the work expectation for adult Danes with children, when the school closures occurred in March, may have been tempered in the short term. As a result, home life with children (at least in the early weeks of the lockdown) may have promoted social interaction and reduced the risk of depression without imposing additional work strain. Future work may want to explicitly consider these important country-level differences when determining what components of the COVID-19 pandemic—the morbidity, the social and educational disruptions, the loss of work—affect changes in mental health and wellbeing. Such work would appear to be critical not only for design of future public health efforts to enhance resilience and recovery, but also for development of theory concerned with collective behavioral responses to adversity.

## Data Availability

Data may be obtained from a third party and are not publicly available. The data used in this study have been made available through a trusted third party, Statistics Denmark. Due to privacy concerns, the data cannot be made available outside the hosted research servers at Statistics Denmark. University-based and private Danish scientific organisations can be authorised to work with data within Statistics Denmark. Such organisations can provide access to individual scientists inside and outside of Denmark. Requests for data may be sent to Statistics Denmark: http://www.dst.dk/en/OmDS/organisation/TelefonbogOrg.aspx?kontor=13&tlfbogsort=sektion or the Danish Data Protection Agency: https://www.datatilsynet.dk/english/the-danish-data-protection-agency/contact/. The authors document and make available all code needed to reproduce the findings in the study.

## DECLARATIONS

### Ethics approval and consent to participate

Statistics Denmark anonymizes and de-identifies the data before making it available to researchers. Use of the data for research purposes is allowed under Danish law for individuals affiliated with Danish research institutions without the need for ethical approval of individual studies. The present study received approval from Statistics Denmark under the auspices of data project no. 707676.

### Consent for publication

Not applicable.

### Competing interests

Non declared.

### Funding

This work was funded by the ROCKWOOL Foundation (grant no. 1227) with additional funding from the Swedish Research Council for Health, Working Life and Welfare (Grant no. 2016-07099) (PF). The research was carried out independently of the funders.

### Authors’ contributions

LHA, PF, and TAB conceived of the presented idea. LHA and PF performed the computations. LHA, PF, and TAB verified the statistical methods. LHA, PF and TAB discussed the results and wrote the manuscript. The corresponding author confirms that he had full access to all the data in the study and had final responsibility for the decision to submit for publication.

## Acknowledgements

The authors thank Laust Hvas Mortensen and Sebastian Simonsen for helpful comments on an earlier draft of the manuscript.

---

**Figure A1.**
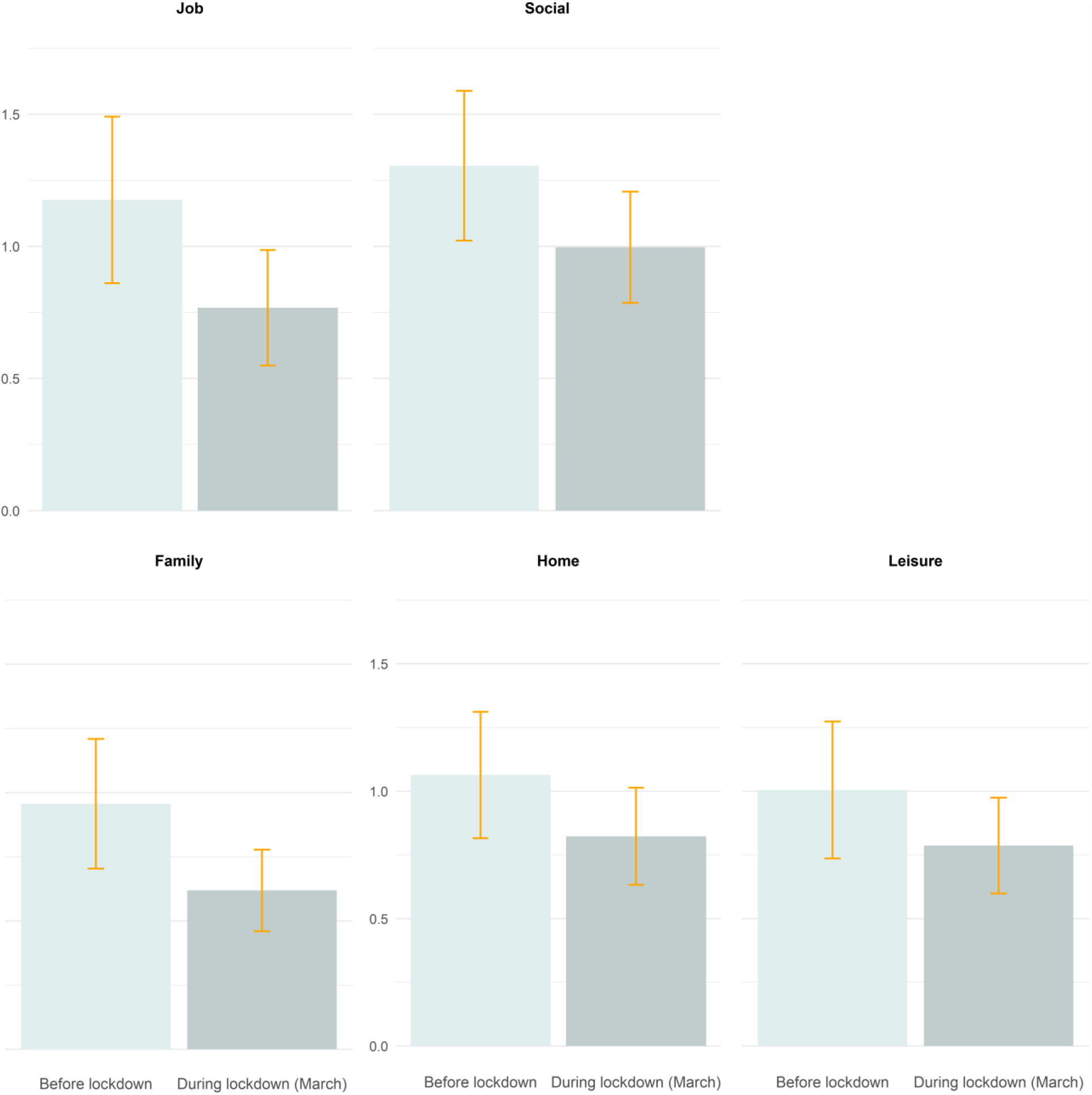
Mean score on subdomains of the WSAS, by time of completing the survey.

**Figure A2.**
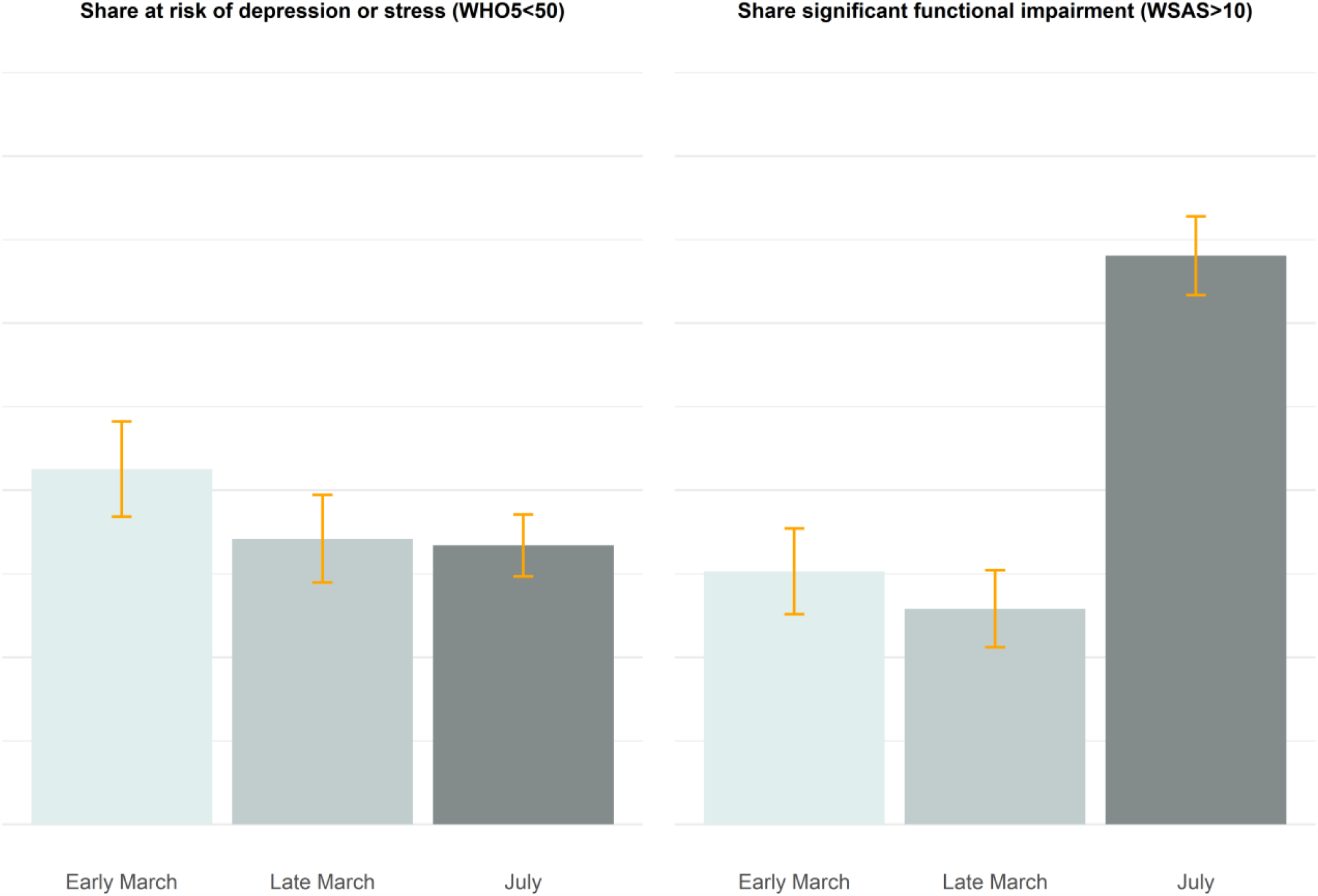
Proportion of respondents at risk of depression or stress according to the WHO5 and proportion of respondents experiencing significant functional impairment according to the WSAS, by time of completing the survey and including respondents in late March.

**Figure A3.**
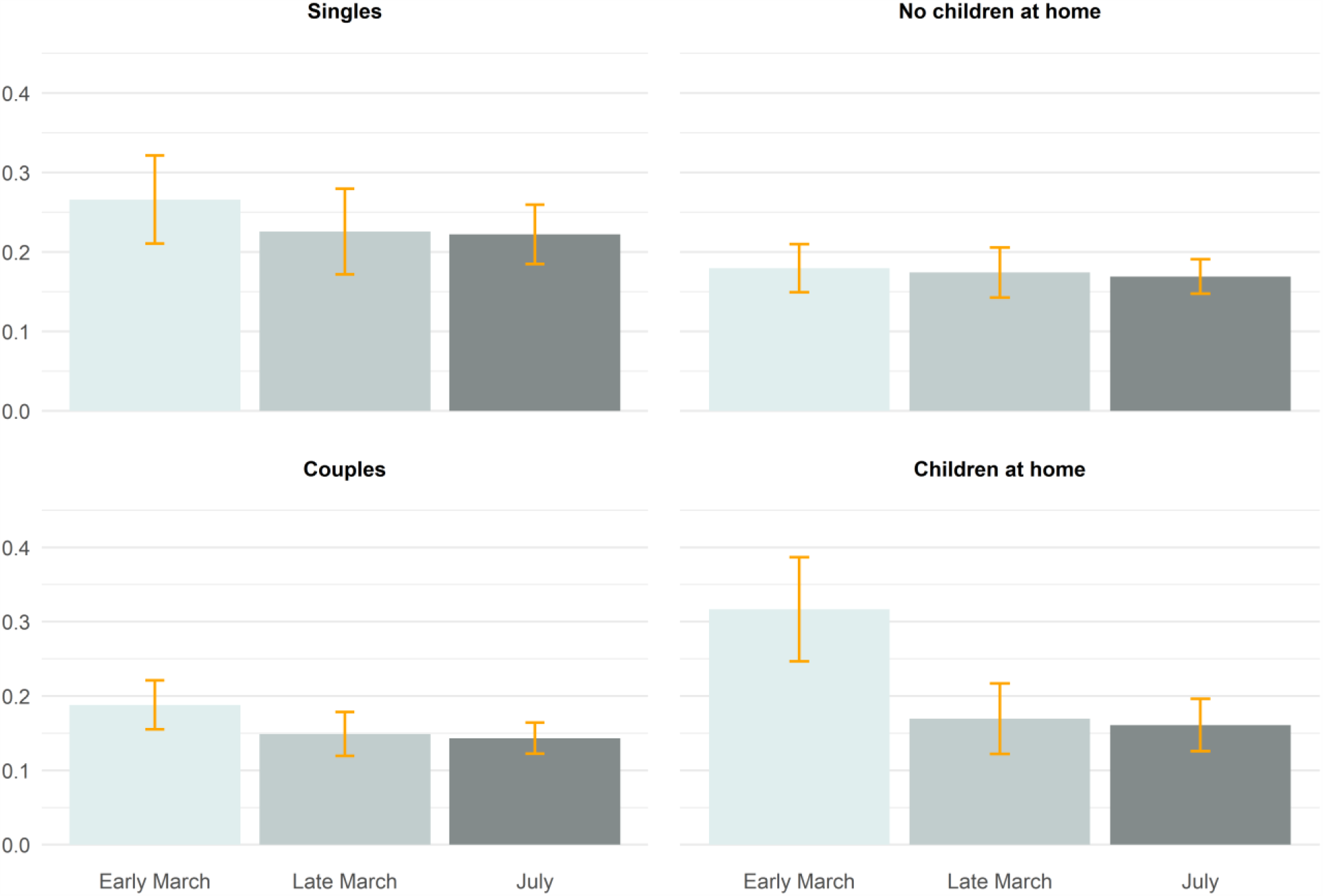
Proportion of respondents at risk of depression or stress according to the WHO5, by time of completing the survey and including respondents in late March.

**Figure A4.**
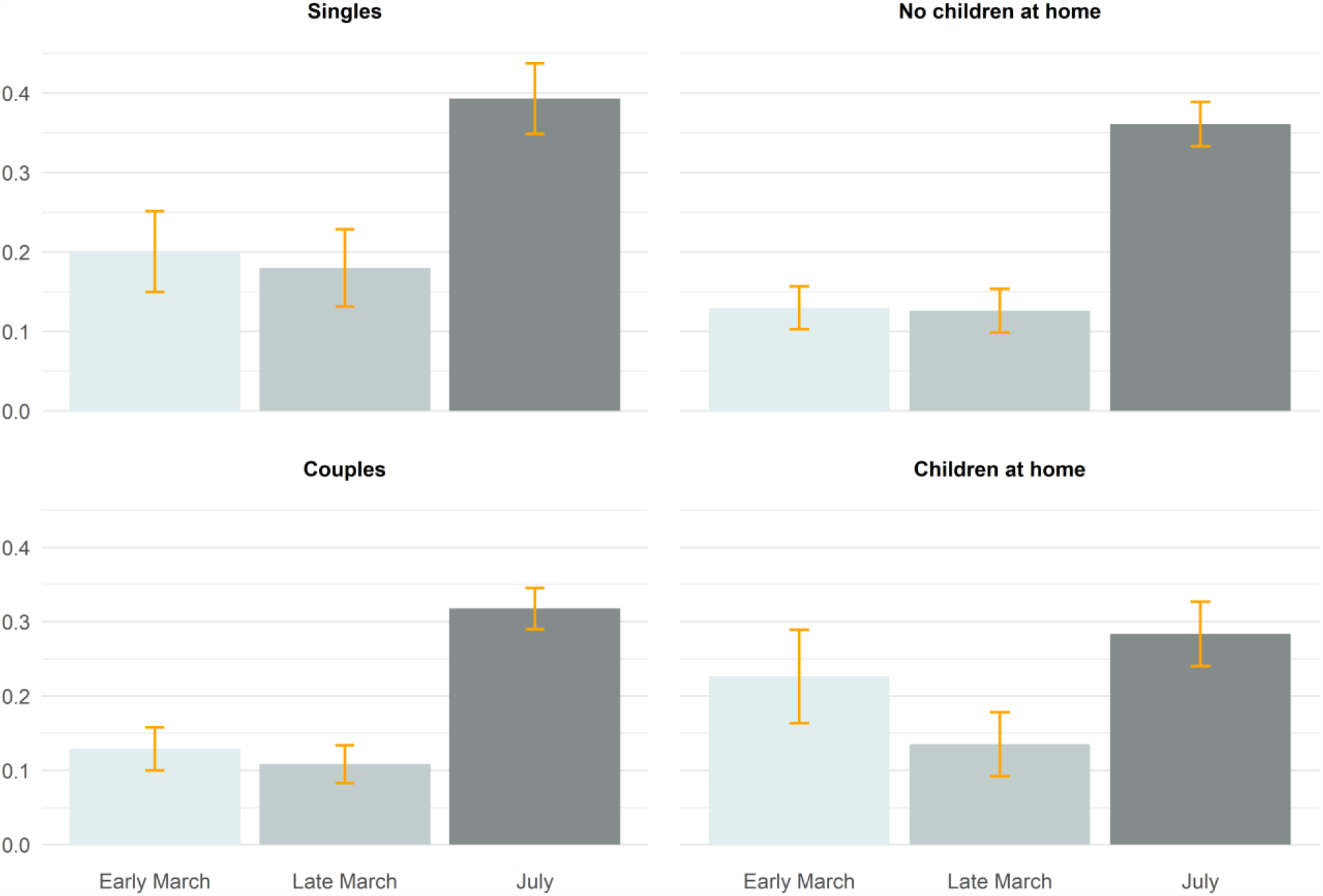
Proportion of respondents with significant functional impairment according to the WSAS, by time of completing the survey and including respondents in late March.

**Figure A5.**
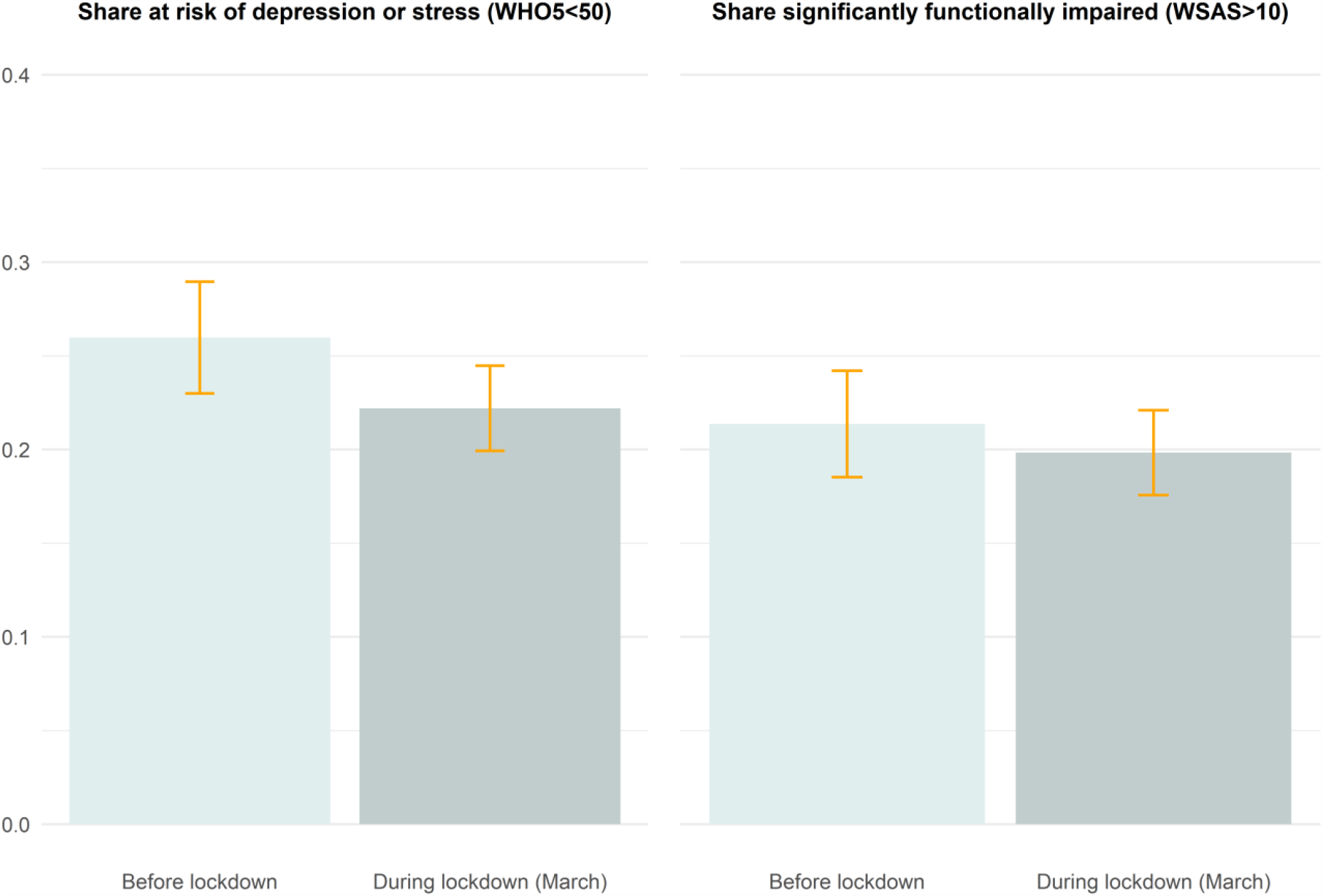
Main results but using population weights from Statistics Denmark instead of statistical controlling.

**Figure A6.**
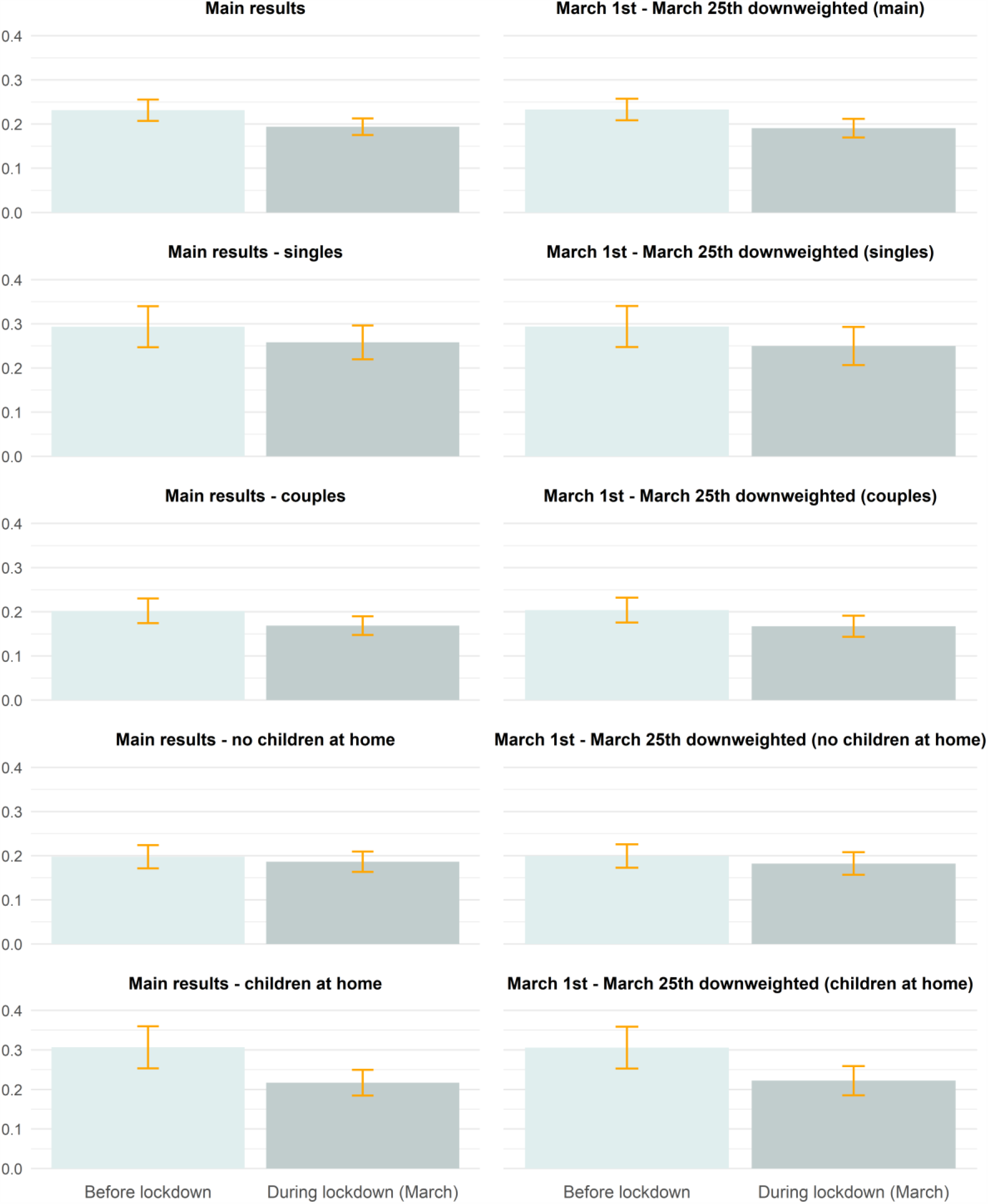
Proportion of respondents at risk of depression or stress according to the WHO5, comparing main results and main results by household structure to results down weighting respondents March 12 – March 25. Note: The down weighting implies weighing individual responses by the numbers of days within the past two weeks prior to survey response that fell on or after the lockdown date, March 11.

**Figure A7.**
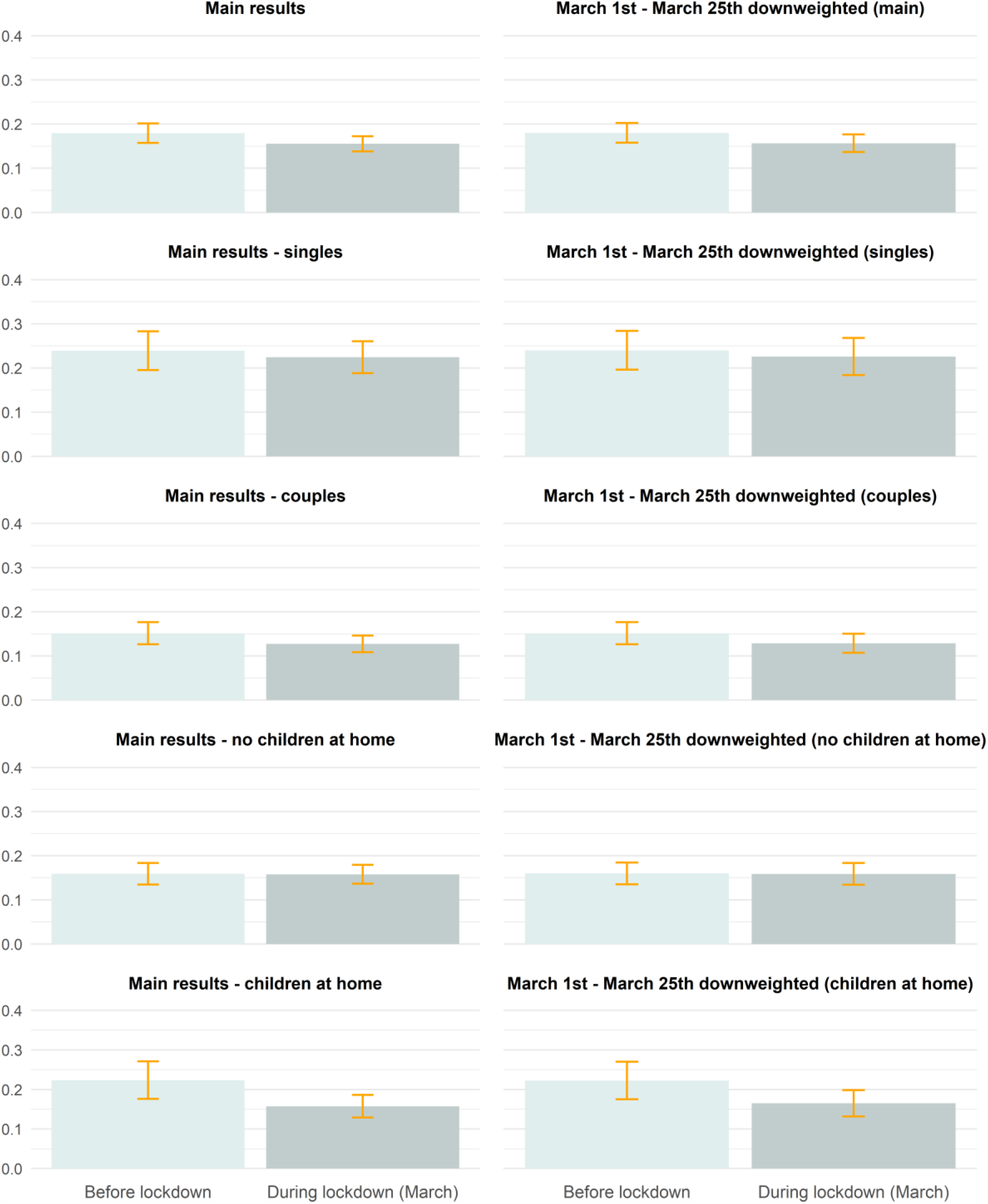
Proportion of respondents significantly functionally impaired according to the WSAS, comparing main results and main results by household structure to results down weighting respondents March 12 – March 25. Note: The down weighting implies weighing individual responses by the numbers of days within the past two weeks prior to survey response that fell on or after the lockdown date, March 11.

**Figure A8.**
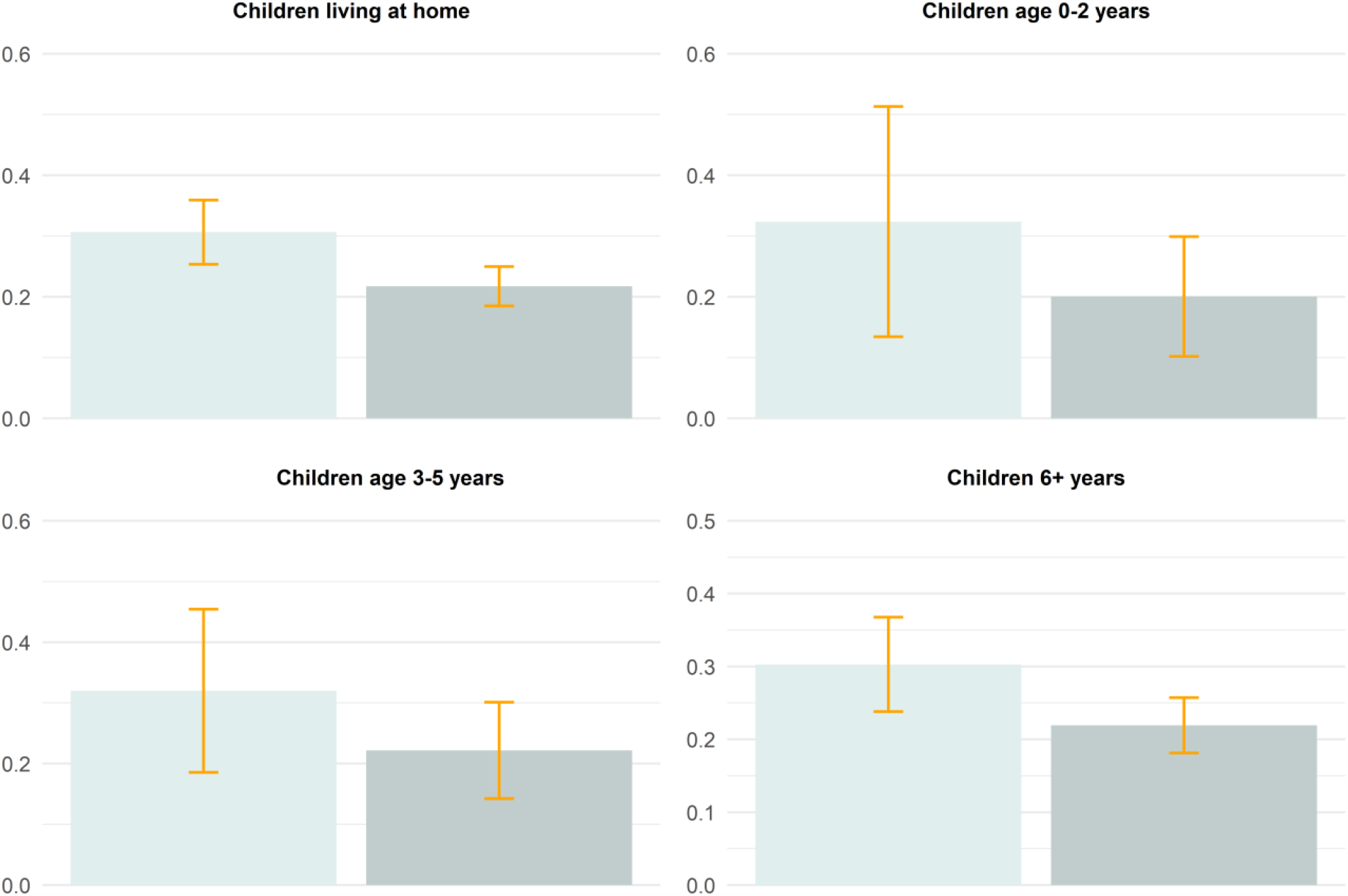
Proportion of respondents at risk of depression or stress according to the WHO5, comparing main results for respondents with children living at home and results by age of the children.

**Figure A9.**
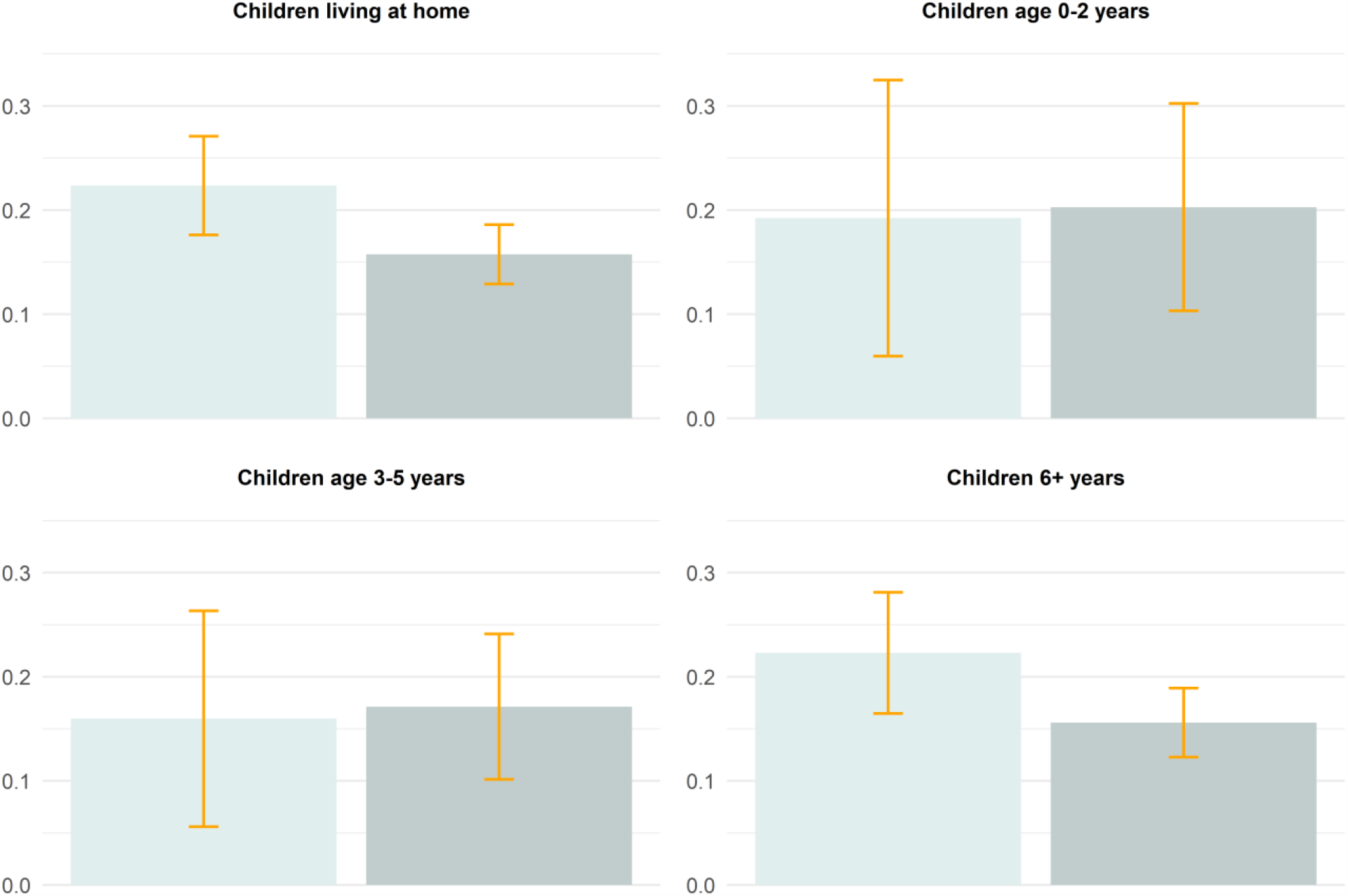
Proportion of respondents significantly functionally impaired according to the WSAS, comparing main results for respondents with children living at home and results by age of the children.

**Table A1.**
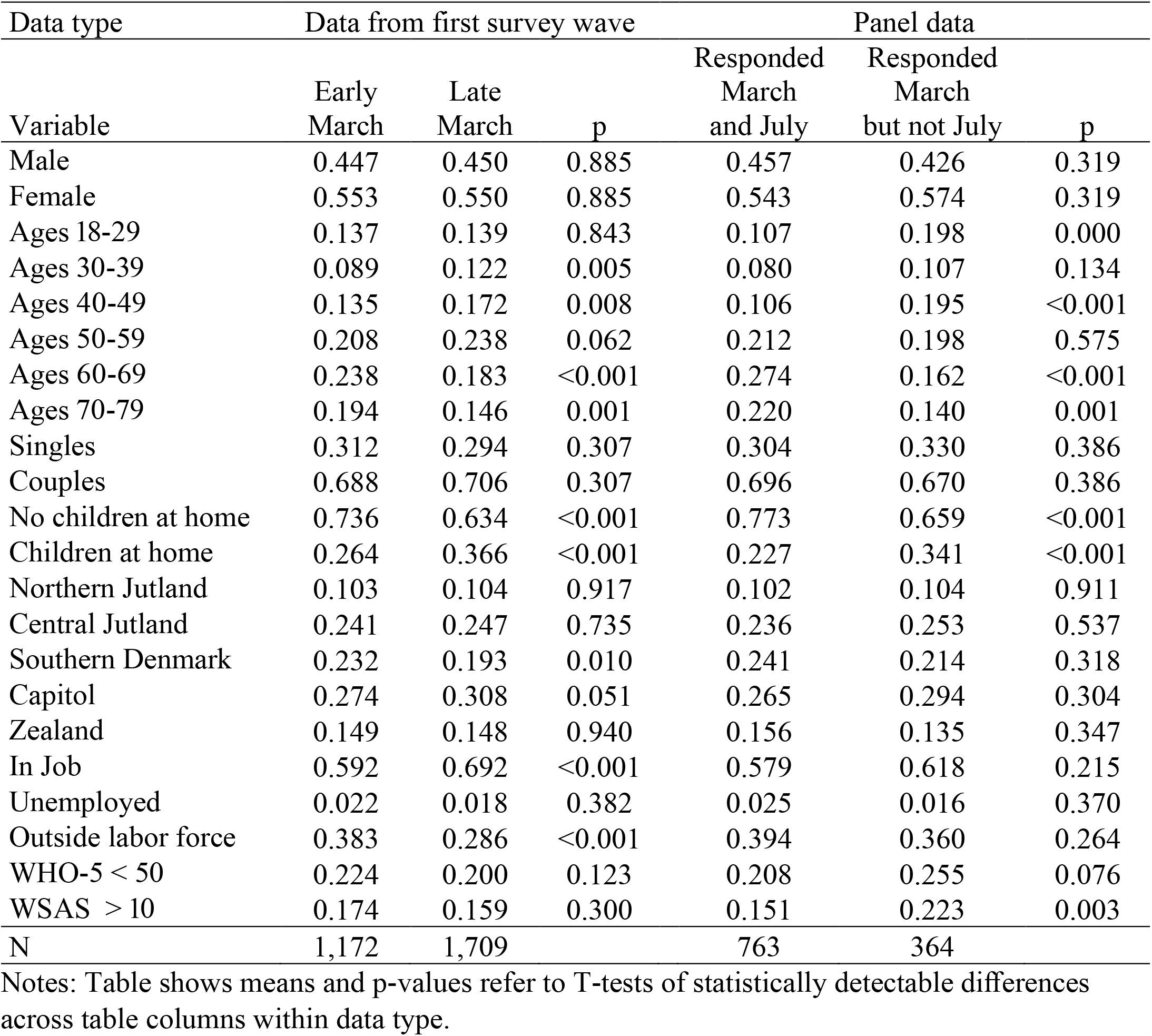
Means of respondents’ background characteristics, by response date and by sample characteristic.

| Data type | Data from first survey wave |  |  | Panel data |  |  |
| --- | --- | --- | --- | --- | --- | --- |
| Variable | Early March | Late March | p | Responded March and July | Responded March but not July | p |
| Male | 0.447 | 0.450 | 0.885 | 0.457 | 0.426 | 0.319 |
| Female | 0.553 | 0.550 | 0.885 | 0.543 | 0.574 | 0.319 |
| Ages 18-29 | 0.137 | 0.139 | 0.843 | 0.107 | 0.198 | 0.000 |
| Ages 30-39 | 0.089 | 0.122 | 0.005 | 0.080 | 0.107 | 0.134 |
| Ages 40-49 | 0.135 | 0.172 | 0.008 | 0.106 | 0.195 | <0.001 |
| Ages 50-59 | 0.208 | 0.238 | 0.062 | 0.212 | 0.198 | 0.575 |
| Ages 60-69 | 0.238 | 0.183 | <0.001 | 0.274 | 0.162 | <0.001 |
| Ages 70-79 | 0.194 | 0.146 | 0.001 | 0.220 | 0.140 | 0.001 |
| Singles | 0.312 | 0.294 | 0.307 | 0.304 | 0.330 | 0.386 |
| Couples | 0.688 | 0.706 | 0.307 | 0.696 | 0.670 | 0.386 |
| No children at home | 0.736 | 0.634 | <0.001 | 0.773 | 0.659 | <0.001 |
| Children at home | 0.264 | 0.366 | <0.001 | 0.227 | 0.341 | <0.001 |
| Northern Jutland | 0.103 | 0.104 | 0.917 | 0.102 | 0.104 | 0.911 |
| Central Jutland | 0.241 | 0.247 | 0.735 | 0.236 | 0.253 | 0.537 |
| Southern Denmark | 0.232 | 0.193 | 0.010 | 0.241 | 0.214 | 0.318 |
| Capitol | 0.274 | 0.308 | 0.051 | 0.265 | 0.294 | 0.304 |
| Zealand | 0.149 | 0.148 | 0.940 | 0.156 | 0.135 | 0.347 |
| In Job | 0.592 | 0.692 | <0.001 | 0.579 | 0.618 | 0.215 |
| Unemployed | 0.022 | 0.018 | 0.382 | 0.025 | 0.016 | 0.370 |
| Outside labor force | 0.383 | 0.286 | <0.001 | 0.394 | 0.360 | 0.264 |
| WHO-5 < 50 | 0.224 | 0.200 | 0.123 | 0.208 | 0.255 | 0.076 |
| WSAS > 10 | 0.174 | 0.159 | 0.300 | 0.151 | 0.223 | 0.003 |
| N | 1,172 | 1,709 |  | 763 | 364 |  |

Notes: Table shows means and p-values refer to T-tests of statistically detectable differences across table columns within data type.

**Table A2.** Parameter estimates from OLS regressions of being at risk of depression/stress (WHO5 < 50) and experiencing significant functional impairment (WSAS > 10).

| Model Outcome | (1)<br>WHO5<50 | (2)<br>WHO5<50 | (3)<br>WSAS>10 | (4)<br>WSAS>10 |
| --- | --- | --- | --- | --- |
| Before 3/11/20 | 0.291***<br>(0.026) | 0.364***<br>(0.038) | 0.183***<br>(0.022) | 0.307***<br>(0.034) |
| After 3/11/20 | -0.037*<br>(0.018) | -0.029<br>(0.016) | -0.024<br>(0.014) | -0.013<br>(0.014) |
| Female | 0.024<br>(0.015) | 0.014<br>(0.015) | 0.022<br>(0.014) | 0.010<br>(0.014) |
| Ages 18-29 | -0.015<br>(0.031) | -0.059<br>(0.032) | 0.072*<br>(0.029) | 0.008<br>(0.029) |
| Ages 30-39 | -0.008<br>(0.033) | -0.014<br>(0.033) | 0.054<br>(0.030) | 0.046<br>(0.029) |
| Ages 50-59 | -0.065*<br>(0.027) | -0.065*<br>(0.028) | -0.012<br>(0.023) | -0.019<br>(0.024) |
| Ages 60-69 | -0.121***<br>(0.026) | -0.160***<br>(0.031) | -0.077***<br>(0.023) | -0.150***<br>(0.027) |
| Ages 70-79 | -0.185***<br>(0.026) | -0.275***<br>(0.033) | -0.076**<br>(0.023) | -0.222***<br>(0.032) |
| Single |  | 0.077***<br>(0.018) |  | 0.070***<br>(0.016) |
| Children at home |  | 0.006<br>(0.022) |  | -0.011<br>(0.019) |
| In Job |  | -0.121***<br>(0.020) |  | -0.181***<br>(0.020) |
| Unemployed |  | -0.071<br>(0.062) |  | -0.171***<br>(0.052) |
| North Jutland |  | 0.006<br>(0.027) |  | 0.046<br>(0.024) |
| Central Jutland |  | -0.004<br>(0.020) |  | 0.041*<br>(0.018) |
| Southern Denmark |  | 0.042<br>(0.022) |  | 0.055**<br>(0.020) |
| Zealand |  | 0.029<br>(0.024) |  | 0.030<br>(0.021) |
| *N* | 2,836 | 2,836 | 2,836 | 2,836 |

Standard errors in parentheses.
\* p < 0.05, \*\* p < 0.01, \*\*\* p < 0.001

**Table A3.** Parameter estimates from OLS regressions of being at risk of depression/stress (WHO5 < 50), by household structure.

| Model<br>Household<br>structure | (1)<br>Singles | (2)<br>Couples | (3)<br>No children at<br>home | (4)<br>Children at home |
| --- | --- | --- | --- | --- |
| Before 3/11/20 | 0.435***<br>(0.053) | 0.239***<br>(0.029) | 0.355***<br>(0.045) | 0.273***<br>(0.039) |
| After 3/11/20 | -0.035<br>(0.031) | -0.034<br>(0.018) | -0.011<br>(0.018) | -0.089**<br>(0.032) |
| Female | -0.031<br>(0.031) | 0.045**<br>(0.017) | 0.009<br>(0.018) | 0.068*<br>(0.029) |
| Ages 18-29 | -0.092<br>(0.057) | -0.014<br>(0.038) | -0.068<br>(0.052) | -0.018<br>(0.043) |
| Ages 30-39 | -0.028<br>(0.069) | -0.011<br>(0.037) | -0.043<br>(0.060) | -0.009<br>(0.039) |
| Ages 50-59 | -0.137*<br>(0.057) | -0.040<br>(0.030) | -0.155**<br>(0.048) | -0.005<br>(0.037) |
| Ages 60-69 | -0.192***<br>(0.056) | -0.096**<br>(0.029) | -0.193***<br>(0.046) | -0.002<br>(0.090) |
| Ages 70-79 | -0.237***<br>(0.057) | -0.166***<br>(0.028) | -0.255***<br>(0.046) | -0.184***<br>(0.029) |
| *N* | 855 | 1981 | 1,913 | 923 |

Standard errors in parentheses
\* $p < 0.05$ , \*\* $p < 0.01$ , \*\*\* $p < 0.001$

**Table A4.** Parameter estimates from OLS regressions being at risk of significant functional impairment (WSAS > 10), by household structure.

| Model<br>Household<br>structure | (1)<br>Singles | (2)<br>Couples | (3)<br>No children at<br>home | (4)<br>Children at home |
| --- | --- | --- | --- | --- |
| Before 3/11/20 | 0.331***<br>(0.051) | 0.131***<br>(0.023) | 0.267***<br>(0.042) | 0.150***<br>(0.033) |
| After 3/11/20 | -0.015<br>(0.029) | -0.024<br>(0.016) | -0.001<br>(0.017) | -0.066*<br>(0.028) |
| Female | -0.029<br>(0.029) | 0.040**<br>(0.015) | 0.009<br>(0.017) | 0.066**<br>(0.025) |
| Ages 18-29 | -0.048<br>(0.054) | 0.106**<br>(0.036) | -0.010<br>(0.049) | 0.091*<br>(0.041) |
| Ages 30-39 | 0.027<br>(0.067) | 0.053<br>(0.033) | 0.021<br>(0.058) | 0.046<br>(0.035) |
| Ages 50-59 | -0.077<br>(0.054) | 0.009<br>(0.025) | -0.111*<br>(0.044) | 0.039<br>(0.031) |
| Ages 60-69 | -0.146**<br>(0.053) | -0.054*<br>(0.023) | -0.170***<br>(0.042) | 0.059<br>(0.083) |
| Ages 70-79 | -0.161**<br>(0.054) | -0.046<br>(0.025) | -0.166***<br>(0.043) | -0.084***<br>(0.024) |
| *N* | 855 | 1,981 | 1,913 | 923 |

Standard errors in parentheses
\* $p < 0.05$ , \*\* $p < 0.01$ , \*\*\* $p < 0.001$

**Table A5.**
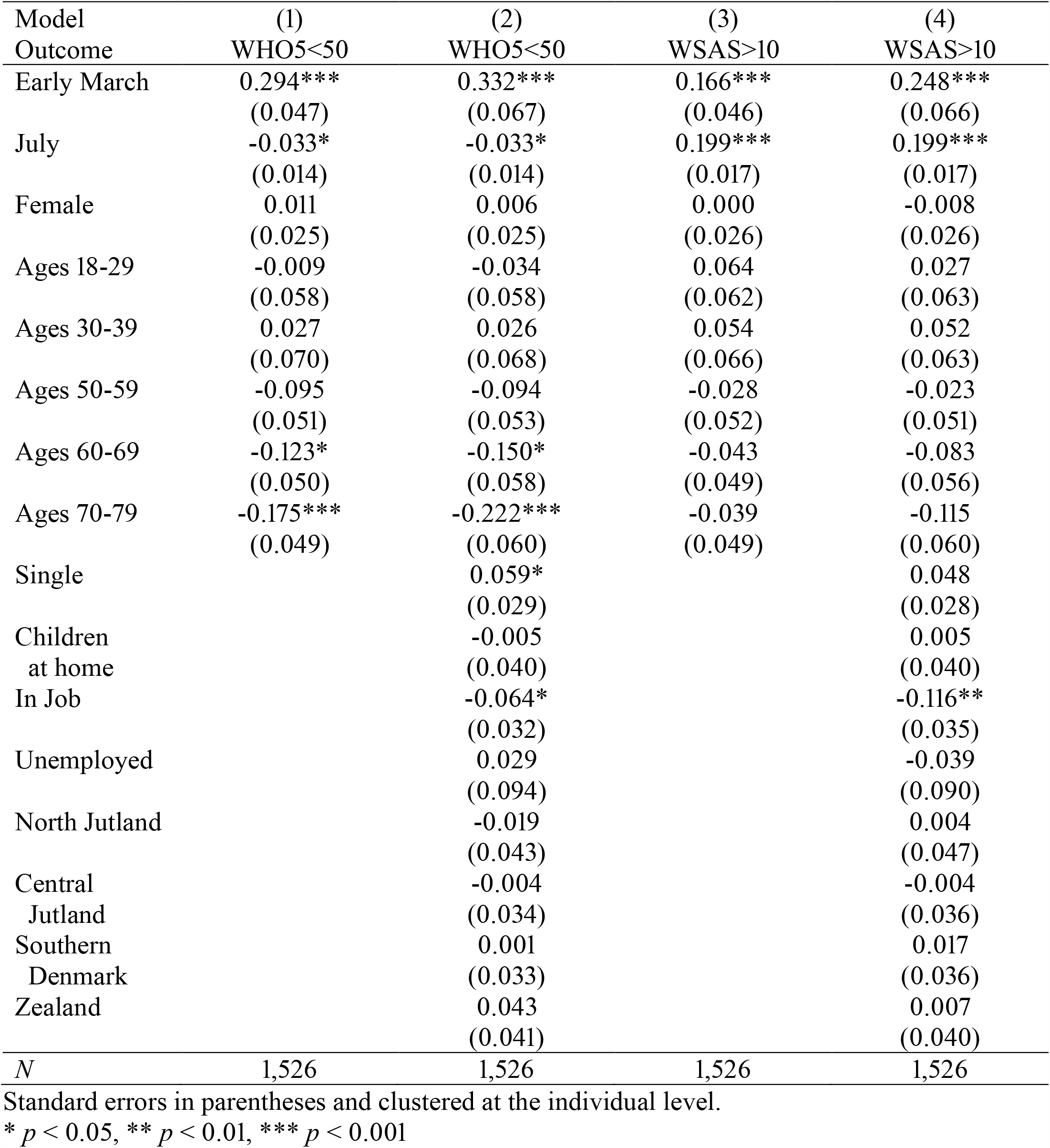
Parameter estimates from OLS regressions of being at risk of depression/stress (WHO5 < 50) and experiencing significant functional impairment (WSAS > 10), repeat responses in early March and July.

**Table A6.**
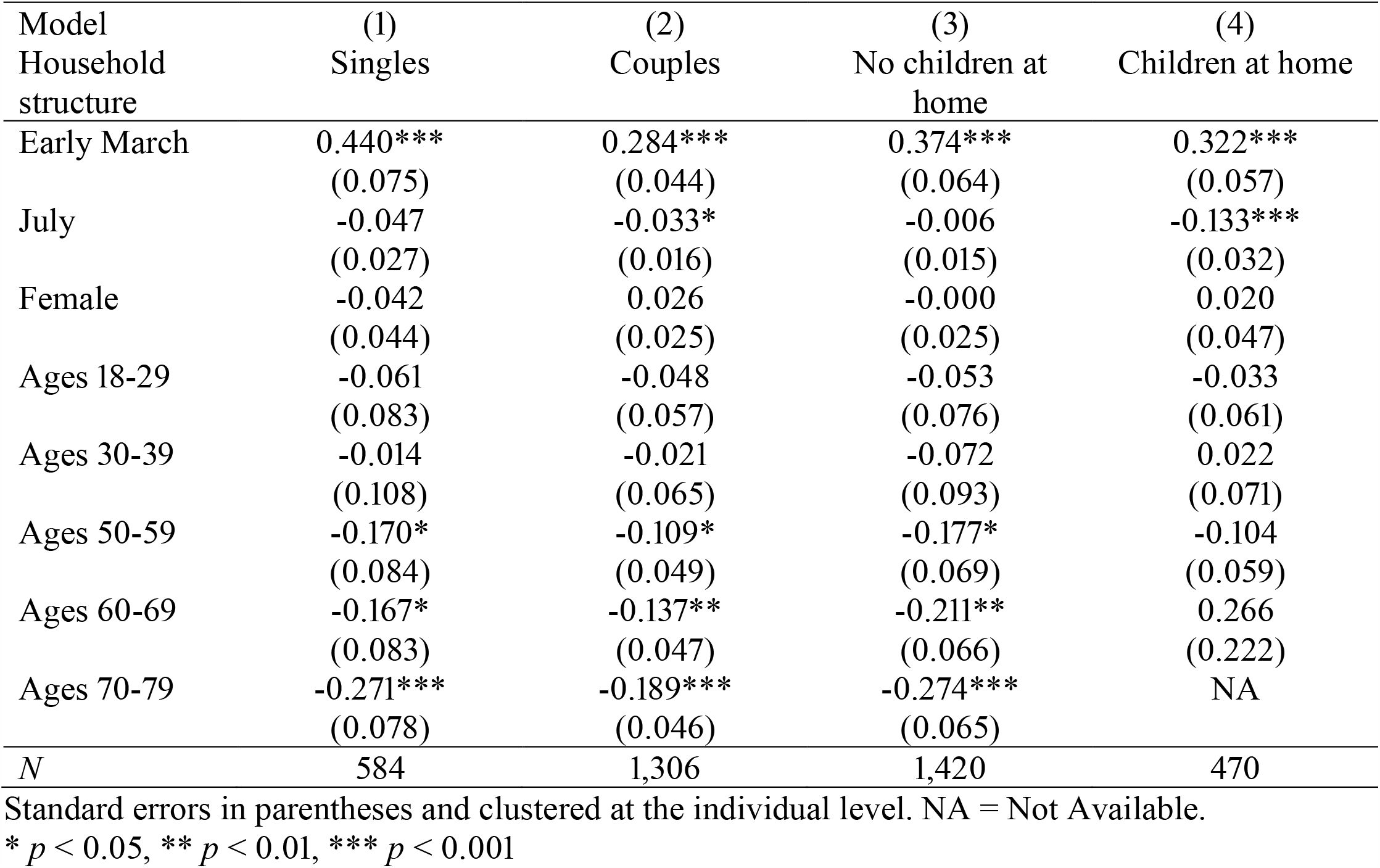
Parameter estimates from OLS regressions of being at risk of depression/stress (WHO5 < 50), by household structure. Repeat responses in early March and July.

**Table A7.** Parameter estimates from OLS regressions of experiencing significant functional impairment (WSAS > 10), by household structure. Repeat responses in early March and July.

| Model<br>Household<br>structure | (1)<br>Singles | (2)<br>Couples | (3)<br>No children at<br>home | (4)<br>Children at home |
| --- | --- | --- | --- | --- |
| Early March | 0.388***<br>(0.075) | 0.117**<br>(0.040) | 0.228***<br>(0.061) | 0.163**<br>(0.050) |
| July | 0.162***<br>(0.033) | 0.191***<br>(0.020) | 0.214***<br>(0.020) | 0.089*<br>(0.035) |
| Female | -0.110*<br>(0.043) | 0.061*<br>(0.026) | -0.016<br>(0.025) | 0.087<br>(0.048) |
| Ages 18-29 | -0.051<br>(0.085) | 0.090<br>(0.063) | 0.055<br>(0.074) | 0.052<br>(0.072) |
| Ages 30-39 | 0.032<br>(0.102) | -0.007<br>(0.059) | 0.057<br>(0.087) | -0.029<br>(0.066) |
| Ages 50-59 | -0.128<br>(0.085) | -0.010<br>(0.047) | -0.092<br>(0.066) | 0.007<br>(0.061) |
| Ages 60-69 | -0.128<br>(0.083) | -0.034<br>(0.044) | -0.099<br>(0.063) | 0.203<br>(0.173) |
| Ages 70-79 | -0.186*<br>(0.078) | -0.007<br>(0.046) | -0.102<br>(0.063) | NA |
| *N* | 584 | 1,306 | 1,420 | 470 |

Standard errors in parentheses and clustered at the individual level. NA = Not Available.
\* $p < 0.05$ , \*\* $p < 0.01$ , \*\*\* $p < 0.001$

**Table A8.**
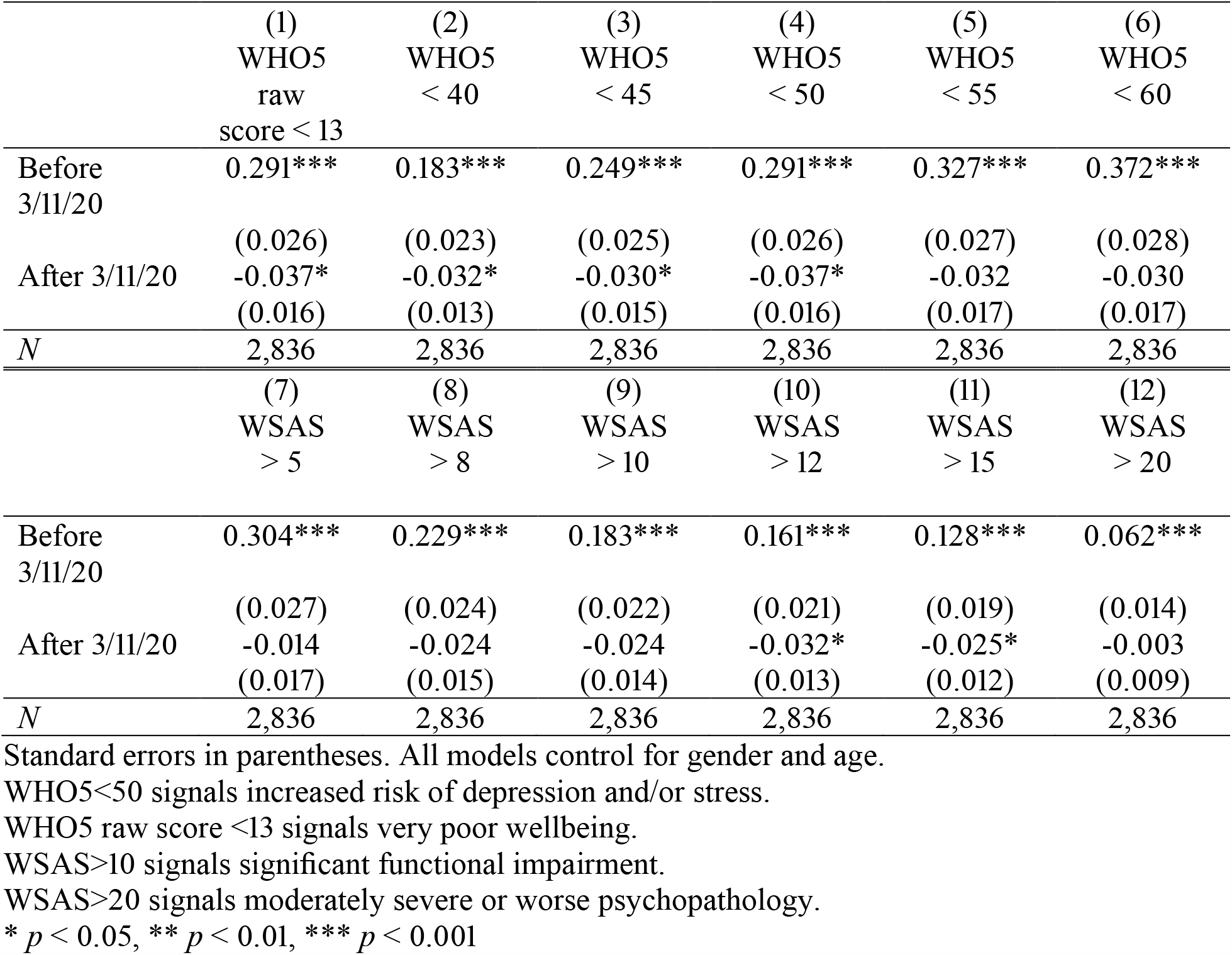
Summary of robustness check of results sensitivity to cutoffs on the WHO5 and WSAS scales.

**Table A9.** Summary of results from individual level fixed effects specification

| WHO-5 < 50 | (1)<br>All | (2)<br>Singles | (3)<br>Couples | (4)<br>No Children<br>at home | (5)<br>Children<br>at home |
| --- | --- | --- | --- | --- | --- |
| Before 3/11/20 | 0.212***<br>(0.012) | 0.275***<br>(0.022) | 0.185***<br>(0.014) | 0.177***<br>(0.012) | 0.324***<br>(0.029) |
| After 3/11/20 | -0.016<br>(0.020) | -0.022<br>(0.038) | -0.014<br>(0.023) | 0.016<br>(0.022) | -0.128**<br>(0.043) |
| Time trend<br>up to July | -0.033*<br>(0.014) | -0.035<br>(0.026) | -0.032*<br>(0.016) | 0.000<br>(0.014) | -0.145***<br>(0.034) |
| *NxT* | 4,393 | 1,323 | 3,070 | 3,054 | 1,339 |
| WSAS > 10 | (6)<br>All | (7)<br>Singles | (8)<br>Couples | (9)<br>No Children<br>at home | (10)<br>Children<br>at home |
| Before 3/11/20 | 0.157***<br>(0.014) | 0.231***<br>(0.026) | 0.126***<br>(0.017) | 0.148***<br>(0.016) | 0.204***<br>(0.029) |
| After 3/11/20 | -0.002<br>(0.023) | -0.026<br>(0.043) | 0.008<br>(0.028) | -0.002<br>(0.028) | -0.041<br>(0.042) |
| Time trend<br>up to July | 0.199***<br>(0.017) | 0.190***<br>(0.032) | 0.203***<br>(0.020) | 0.232***<br>(0.020) | 0.087*<br>(0.034) |
| *NxT* | 4,393 | 1,323 | 3,070 | 3,054 | 1,339 |

Standard errors in parentheses
\* $p < 0.05$ , \*\* $p < 0.01$ , \*\*\* $p < 0.001$

## Notes

### Competing Interest Statement

The authors have declared no competing interest.

